# Factors associated with self-reported anxiety, depression, and general health during the UK lockdown; a cross-sectional survey

**DOI:** 10.1101/2020.06.23.20137901

**Authors:** Louise E. Smith, Richard Amlot, Helen Lambert, Isabel Oliver, Charlotte Robin, Lucy Yardley, G James Rubin

## Abstract

**Background:** To investigate factors associated with anxiety, depression, and self-reported general health during “lockdown” due to COVID-19 in the UK.

**Methods:** Online cross-sectional survey of a nationally-representative sample of 2240 participants living in the UK aged 18 years or over (data collected 6-7 May 2020). Participants were recruited from YouGov’s online research panel.

**Outcomes:** In this sample, 21·9% (n=458, 95% CI [20·1% to 23·7%]) reported probable anxiety (scored three or over on the GAD-2); while 23·5% (n=494, 95% CI [21·7% to 25·3]) reported probable depression (scored three or over on the PHQ-2). Poorer mental health was associated with greater financial hardship during the lockdown, thinking that you would lose contact with friends or family if you followed Government measures, more conflict with household members during the lockdown, less sense of community with people in your neighbourhood, and lower perceived effectiveness of Government measures. Females and those who were younger were likely to report higher levels of anxiety and depression. The majority of participants reported their general health as “good” (as measured by the first item of the SF-36). Poorer self-reported general health was associated with psychological distress, greater worry about COVID-19 and markers of inequality.

**Interpretation:** Rates of self-reported anxiety and depression in the UK during the lockdown were greater than population norms. Reducing financial hardship, promoting social connectedness, and increasing solidarity with neighbours and household members may help ease rifts within the community which are associated with distress, thereby improving mental health. Reducing inequality may also improve general health.

**RESEARCH IN CONTEXT:** *Evidence before this study:*

- Quarantine is associated with adverse psychological outcomes.
- Psychological distress during quarantine is associated with greater financial loss, greater perceived susceptibility to and severity of the illness, and greater frustration and boredom during quarantine.
- Measures put in place to prevent the spread of COVID-19 have highlighted existing inequalities in society, disproportionally affecting younger people, those in lower-income households, and Black and minority ethnic groups.
- Research in the UK and other countries indicates that rates of anxiety and depression during restrictions of movement such as “lockdown” measures are higher than population norms.

*Added value of this study:*

- In this study, 22% of the sample reported anxiety, while 24% reported depression. Normative data indicate that these rates are usually approximately 5% and 7% respectively.
- Factors associated with psychological distress included greater financial hardship, poorer social connectedness, greater conflict within the household and the wider neighbourhood, being female and of younger age.
- Self-reported general health in the sample was “good” on average. Factors associated with poorer self-reported general health included markers of inequality and greater worry about COVID-19.

*Implications of all the available evidence:*

- Decreasing the financial impact of measures put in place to prevent the spread of COVID-19 may help improve mental health.
- Interventions promoting social connectedness in isolated young people and measures that increase household and neighbourhood solidarity may help improve mental health.

## INTRODUCTION

The COVID-19 pandemic has seen unprecedented levels of restrictions of movement worldwide. Quarantine has negative psychological consequences.^1^ Psychological distress from quarantine is associated with situational factors such as longer duration of quarantine, having inadequate supplies (such as food, clothes, or accommodation) and greater financial loss, and psychological factors, such as greater perceived susceptibility to and severity of the illness, frustration, boredom, inadequate information, and stigmatisation.^1^ The UK Government imposed “lockdown” measures restricting people’s movement apart from for limited, specified reasons on 23^rd^ March 2020,^2^ which were slightly eased on 11^th^ May 2020.^3^ The pandemic has highlighted inequalities already present in society. For example, evidence shows that measures put in place to prevent the spread of COVID-19 have disproportionally affected females, younger people, those in low-income households and Black and minority ethnic groups.^4^

Research is beginning to emerge indicating that those under lockdown measures have higher levels of psychological distress.^5–8^ In the UK, preliminary research has shown that average levels of anxiety, depression and stress were higher during the lockdown than population norms.^9^ However, these results should be taken with caution as the sample was not demographically representative of the UK population (e.g. 85% female) and data were collected across almost four weeks. Results from a demographically-representative sample of the UK population show that anxiety was associated with loneliness, female sex, and one’s work being affected by the COVID-19 pandemic.^10^ However, anxiety was not measured using a validated scale. While reported levels of mental distress should be taken with caution in rapidly constructed samples,^11^ where studies are rigorously conducted, associations within the data may still provide useful insights.^12^ Rapid research undertaken during public health crises often results in decreased methodological rigour.^12,13^ Online quota sampling is a standard approach used in opinion polling and can be a pragmatic approach when a large, demographically representative sample must be obtained in a very short time frame, particularly during a crisis.^12^

In this study, we investigated psychological, situational, personal and clinical factors associated with validated measures of anxiety and depression, and self-reported general health in a demographically representative sample of the UK adult population during lockdown.

## METHOD

### Design

We commissioned YouGov Plc, a market research company, to carry out this cross-sectional survey, between 6^th^ and 7^th^ May 2020.

Results from analyses investigating adherence to self-isolation and lockdown measures using the same sample have been reported elsewhere.^14,15^

### Participants

Participants (n=2240) were recruited from YouGov’s online research panel (n=800,000+ UK adults) and were eligible for the study if they were aged eighteen years or over and lived in the UK. To ensure that the sample was broadly representative of the UK general population, we used quota sampling. Quotas were set based on age, gender, social grade and highest level of education, and Government Office Region. Of 2,623 people who started the survey, 2,314 completed it. Seventy-four people were excluded from the sample as data were missing for demographic questions, or their response did not meet quality control standards (completed the survey suspiciously quickly or gave identical answers for multiple consecutive questions). Participants who completed the survey were reimbursed in points equivalent to approximately 50 pence.

### Study materials

Full survey materials are available in the supplementary materials.

#### Outcome measures

We measured probable anxiety using the validated GAD-2.^16^ We measured probable depression using the validated PHQ-2.^17^ For these items, participants were asked to rate how much they had been bothered in the past two weeks on a four-point Likert-type scale from “not at all” to “nearly every day”. Participants could also answer “don’t know” and “prefer not to say” to these questions.

Participants rated their general health on a five-point Likert-type scale from “poor” to “excellent” using the first question from the SF-36.^18^

#### Psychological and situational factors

We asked participants whether they thought they “had, or currently have, coronavirus”. Possible answers were “I have definitely had it or definitely have it now”, “I have probably had it or probably have it now”, “I have probably not had it and probably don’t have it now”, and “I have definitely not had it and definitely don’t have it now”.

Participants reported whether they had experienced symptoms “in the past seven days” from a list of thirteen symptoms including cough and high temperature / fever. Participants who lived with someone else also reported if “someone else in [their] household” had experienced symptoms “in the past fourteen days” from the same list of thirteen symptoms.

Participants were asked to indicate whether they were currently self-isolating, choosing between “not self-isolating”, “self-isolating for seven days”, “self-isolating for fourteen days”, and “self-isolating for at least 12 weeks”.

Participants reported “in the past seven days, how many times, if at all, [they had] left [their] home” to go to the shops for groceries, toiletries or medicine, to go to the shops for other items, for exercise, for a medical purpose, to go to work, and to help someone else.

We asked participants how worried they were about COVID-19 on a five-point Likert-type scale from “not at all worried” to “extremely worried”.

Participants were asked approximately what percentage of people of around the same age they thought were “fully following the UK Government’s recommendations to stay at home”.

Participants reported whether they thought the current lockdown had made their physical health better or worse. Possible answers were “a lot better”, “a little better”, “no difference”, “a little worse”, and “a lot worse”.

Participants were asked if they had helped, or received help from, someone outside their household in the past seven days (yes / no).

Participants rated fourteen perception statements on a five-point Likert scale from “strongly disagree” to “strongly agree”. Statements included perceived severity of COVID-19, perceived effectiveness of Government measures, perceived likelihood of catching and spreading COVID-19, perceived costs of following Government measures, fear of losing touch with friends and relatives, social pressure from friends and family to follow Government measures, perceived legal consequences from not following Government measures, and positive consequences of the lockdown.

#### Personal and clinical characteristics

We asked participants to report their age, gender, employment status, highest educational or professional qualification, and their marital status. We also asked participants whether there was a child in their household, whether they or someone else in their household received a letter from the NHS telling them they were extremely clinically vulnerable to COVID-19 (proxy for chronic illness), and whether they lived alone. Participants were asked for their postcode to determine indices of multiple deprivation and whether they lived in an urban or rural area. We also collected social grade.

Participants were asked if their primary home had access to any outdoor space, and whether they were pet owners.

### Ethics

Ethical approval for this study was granted by the King’s College London Research Ethics Committee (reference: LRS-19/20-18687).

### Patient and public involvement

Due to the rapid nature of this research, the public was not involved in the development of the survey materials.

### Power

A sample size of 2,240 allows a 95% confidence interval of plus or minus 2% for the prevalence estimate for each survey item.

### Analysis

#### Recoding variables

GAD-2 scores of three or over indicated probable anxiety.^16^ PHQ-2 scores of three or over indicated probable depression.^17^ We were unable to calculate a score for anxiety or depression for people who skipped, or answered “don’t know” or “prefer not to say”, to one or both questions for these scales (GAD-2: 6.0% of the sample, n=134. PHQ-2: 5.4% of the sample, n=122).

Presence of COVID-19 symptoms in the household was recoded into a binary variable. We defined presence of COVID-19 symptoms as reporting having cough or high temperature / fever oneself in the last seven days, or reporting that a household member had a cough or high temperature / fever in the last fourteen days.

We recoded whether participants reported self-isolating into a binary variable (not self-isolating, self-isolating).

We calculated the total number of outings made in the last seven days by summing the number of times people reported going out (shopping for groceries, toiletries or medicine, shopping for other items, exercising, for a medical purpose, going to work, helping someone else, meeting friends or family). We grouped participants who reported going out more than twenty times in the past seven days (n=54, 2.4%).

For all variables, unless stated otherwise, we coded answers of “don’t know” or “prefer not to say” as missing data.

#### Analyses

We ran a series of logistic regressions investigating univariable associations between personal and clinical factors, psychological and situational factors and anxiety and depression. We ran a second set of logistic regressions controlling for personal and clinical characteristics.

We ran a series of linear regressions investigating univariable associations between personal and clinical factors, psychological and situational factors and self-rated general health. We ran a second set of linear regressions controlling for personal and clinical characteristics. For these regressions, we used the total proportion of the variance explained by individual psychological and situational factors, in addition to personal and clinical characteristics, and statistical significance of individual regression coefficients to determine which factors were most strongly associated.

Data were weighted to increase their representativeness of the UK population.

#### Sensitivity analyses

As we ran many analyses on each outcome (n=39 for anxiety and depression, n=40 for self-reported general health), we applied a Bonferroni correction to our results (*p*≤·001). Analyses reaching this significance level are marked by a double asterisk (**) in the tables.

## RESULTS

Only results of adjusted analyses are reported narratively. Results of unadjusted analyses are reported in the tables.

### Anxiety

Data were available to calculate anxiety scores for 2,089 participants. 21·9% (n=458, 95% CI [20·1% to 23·7%]) reported probable anxiety (scored three or over on GAD-2).

Anxiety was associated with being female, younger, not working, and living in a more deprived area (Table 1).

**Table 1.** Associations between personal and clinical characteristics and anxiety and depression.

| Participant characteristics | Level | Psychological distress |  | Odds ratio (95% CI) | Adjusted odds ratio (95% CI)† | No probable depression n=1610, n (%) | Probable depression n=494, n (%) | Odds ratio (95% CI) | Adjusted odds ratio (95% CI)† |
| --- | --- | --- | --- | --- | --- | --- | --- | --- | --- |
|  |  | No probable anxiety n=1632, n (%) | Probable anxiety n=458, n (%) |  |  |  |  |  |  |
| Gender | Male | 829 (82.8) | 172 (17.2) | Reference | Reference | 796 (78.0) | 225 (22.0) | Reference | Reference |
|  | Female | 803 (73.8) | 285 (26.2) | 1.71 (1.38 to 2.12)** | 2.00 (1.57 to 2.55)** | 814 (75.2) | 269 (24.8) | 1.17 (0.96 to 1.43) | 1.23 (0.98 to 1.54) |
| Age | 18 to 24 years | 126 (58.6) | 89 (41.4) | Reference | Reference | 130 (58.6) | 92 (41.4) | Reference | Reference |
|  | 25 to 34 years | 219 (72.3) | 84 (27.7) | 0.55 (0.38 to 0.79)** | 0.50 (0.31 to 0.79)* | 213 (71.2) | 86 (28.8) | 0.57 (0.40 to 0.83)* | 0.66 (0.41 to 1.04) |
|  | 35 to 44 years | 298 (79.3) | 78 (20.7) | 0.37 (0.26 to 0.54)** | 0.30 (0.19 to 0.48)** | 278 (74.9) | 93 (25.1) | 0.48 (0.33 to 0.68)** | 0.55 (0.35 to 0.87)* |
|  | 45 to 54 years | 274 (77.0) | 82 (23.0) | 0.42 (0.29 to 0.61)** | 0.37 (0.23 to 0.59)** | 268 (75.3) | 88 (24.7) | 0.47 (0.33 to 0.67)** | 0.56 (0.36 to 0.90)* |
|  | 55 years and over | 715 (85.1) | 125 (14.9) | 0.25 (0.18 to 0.35)** | 0.21 (0.13 to 0.33)** | 722 (84.3) | 134 (15.7) | 0.26 (0.19 to 0.36)** | 0.26 (0.17 to 0.41)** |
| Child in the household | None | 1201 (79.5) | 309 (20.5) | Reference | Reference | 1181 (77.6) | 341 (22.4) | Reference | Reference |
|  | Present | 415 (75.5) | 135 (24.5) | 1.27 (1.01 to 1.60)* | 1.01 (0.76 to 1.35) | 411 (74.6) | 140 (25.4) | 1.18 (0.94 to 1.48) | 1.06 (0.80 to 1.40) |
| Clinically extremely vulnerable (self) | No | 1491 (78.5) | 409 (21.5) | Reference | Reference | 1481 (77.7) | 424 (22.3) | Reference | Reference |
|  | Yes | 99 (77.3) | 29 (22.7) | 1.07 (0.70 to 1.64) | 1.18 (0.74 to 1.9) | 92 (70.2) | 39 (29.8) | 1.49 (1.01 to 2.20)* | 1.70 (1.11 to 2.59)* |
| Employment status | Not working | 741 (77.0) | 221 (23.0) | Reference | Reference | 733 (75.4) | 239 (24.6) | Reference | Reference |
|  | Working | 891 (79.1) | 236 (20.9) | 0.89 (0.72 to 1.09) | 0.71 (0.54 to 0.92)* | 877 (77.5) | 254 (22.5) | 0.89 (0.73 to 1.09) | 0.65 (0.50 to 0.85)** |
| Highest educational or professional qualification | GCSE/vocational/A-level/No formal qualifications | 755 (77.0) | 225 (23.0) | Reference | Reference | 748 (75.5) | 243 (24.5) | Reference | Reference |
|  | Degree or higher (Bachelors, Masters, PhD) | 856 (79.7) | 218 (20.3) | 0.86 (0.69 to 1.06) | 0.90 (0.70 to 1.14) | 841 (78.1) | 236 (21.9) | 0.86 (0.70 to 1.06) | 1.00 (0.79 to 1.27) |
| IMD decile | Range 1 (most deprived) to 10 (least deprived) | N=1632, M=6.06, SD=2.75 | N=458, M=5.31, SD=2.93 | 0.91 (0.88 to 0.94)** | 0.92 (0.88 to 0.96)** | N=1610, M=6.11, SD=2.93 | N=494, M=5.32, SD=2.92 | 0.90 (0.87 to 0.94)** | 0.93 (0.89 to 0.97)** |
| Social grade | ABC1 | 950 (79.4) | 247 (20.6) | Reference | Reference | 940 (78.0) | 265 (22.0) | Reference | Reference |
|  | C2DE | 682 (76.4) | 211 (23.6) | 1.19 (0.97 to 1.47) | 1.04 (0.81 to 1.33) | 670 (74.5) | 229 (25.5) | 1.22 (0.99 to 1.49) | 1.12 (0.88 to 1.42) |
| Urban/rural | Urban | 1233 (76.7) | 374 (23.3) | Reference | Reference | 1206 (74.7) | 408 (25.3) | Reference | Reference |
|  | Rural | 360 (84.1) | 68 (15.9) | 0.62 (0.47 to 0.82)** | 0.79 (0.57 to 1.08) | 361 (83.2) | 73 (16.8) | 0.60 (0.45 to 0.79)** | 0.75 (0.55 to 1.02) |
| Living alone | Yes | 319 (79.9) | 80 (20.1) | Reference | Reference | 294 (72.1) | 114 (27.9) | Reference | Reference |
|  | No | 1313 (77.8) | 375 (22.2) | 1.14 (0.87 to 1.50) | 1.04 (0.70 to 1.53) | 1316(77.6) | 379 (22.4) | 0.74 (0.58 to 0.95)* | 0.73 (0.51 to 1.04) |
| Marital status | Married/civil partnership/living as married | 996 (80.8) | 236 (19.2) | Reference | Reference | 1003 (81.6) | 226 (18.4) | Reference | Reference |
|  | Separated/divorced/widowed/never married | 626 (74.1) | 219 (25.9) | 1.48 (1.20 to 1.82)** | 1.07 (0.78 to 1.48) | 594 (69.2) | 264 (30.8) | 1.97 (1.61 to 2.42)** | 1.31 (0.96 to 1.80) |
| Clinically extremely vulnerable (household member) | No | 1171 (78.4) | 323 (21.6) | Reference | Reference | 1165 (78.2) | 324 (21.8) | Reference | Reference |
|  | Yes | 111 (75.5) | 36 (24.5) | 1.18 (0.79 to 1.75) | 1.01 (0.64 to 1.59) | 124 (80.0) | 31 (20.0) | 0.89 (0.59 to 1.34) | 0.61 (0.37 to 1.00)* |
| Home includes access to outside space | No | 128 (74.0) | 45 (26.0) | Reference | Reference | 113 (68.5) | 52 (31.5) | Reference | Reference |
|  | Yes | 1504 (78.5) | 412 (21.5) | 0.78 (0.54 to 1.11) | 1.03 (0.66 to 1.60) | 1497 (77.2) | 441 (22.8) | 0.64 (0.45 to 0.90)* | 0.93 (0.61 to 1.41) |
| Pet ownership | No | 878 (78.5) | 241 (21.5) | Reference | Reference | 878 (78.0) | 247 (22.0) | Reference | Reference |
|  | Yes | 754 (77.7) | 216 (22.3) | 1.04 (0.85 to 1.29) | 0.95 (0.75 to 1.22) | 731 (74.7) | 247 (25.3) | 1.20 (0.98 to 1.47) | 1.16 (0.91 to 1.46) |

\* $p \leq .05$
\*\* $p \leq .001$
† Adjusting for gender, age, having a child in the household, being extremely clinically vulnerable oneself, employment status, highest level of education or professional qualification, indices of multiple deprivation, social grade, living in a rural or urban area, living alone, marital status, and region.
‡ Adjusted analyses for this variable did not control for living alone, as by definition all participants asked this question lived in a household with someone else.

Anxiety was also associated with: the presence of depression; feeling that the current lockdown had made your mental health worse; presence of COVID-19 symptoms in the household; greater worry about COVID-19; having received help from someone outside your household because of COVID-19 in the last week; having more conflict in your household because of the lockdown; reporting that you were currently self-isolating; not enjoying spending more time at home during the lockdown; believing that following Government advice would impact your finances negatively and cause you to lose touch with friends and relatives; greater perceived severity of COVID-19 for your family’s wellbeing; lower perceived sense of community with people in your neighbourhood; lower perceived effectiveness of Government measures; and thinking that fewer people of your age in the UK were following the UK Government’s recommendations to stay at home (lower perceived social norms; Table 2).

**Table 2.**
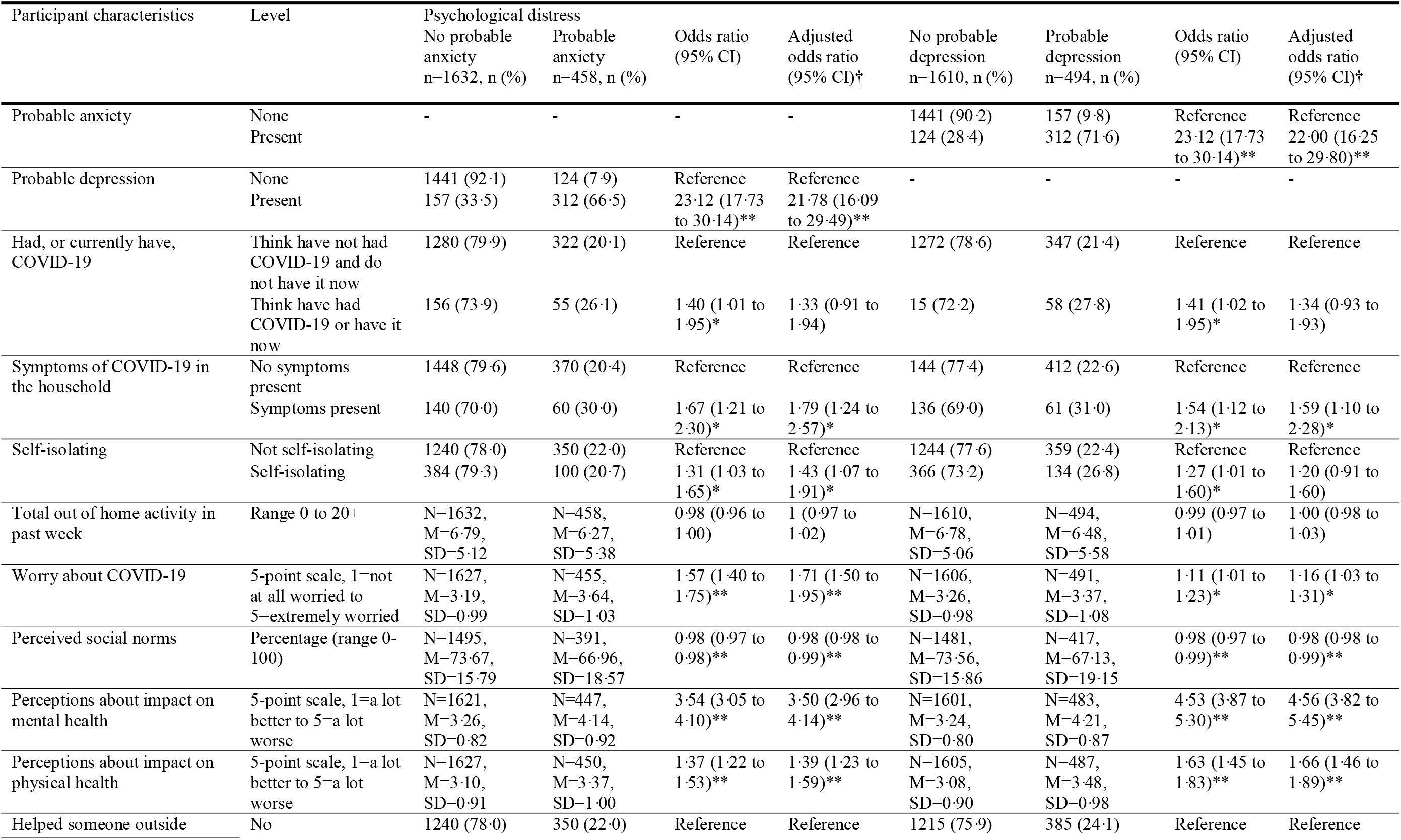
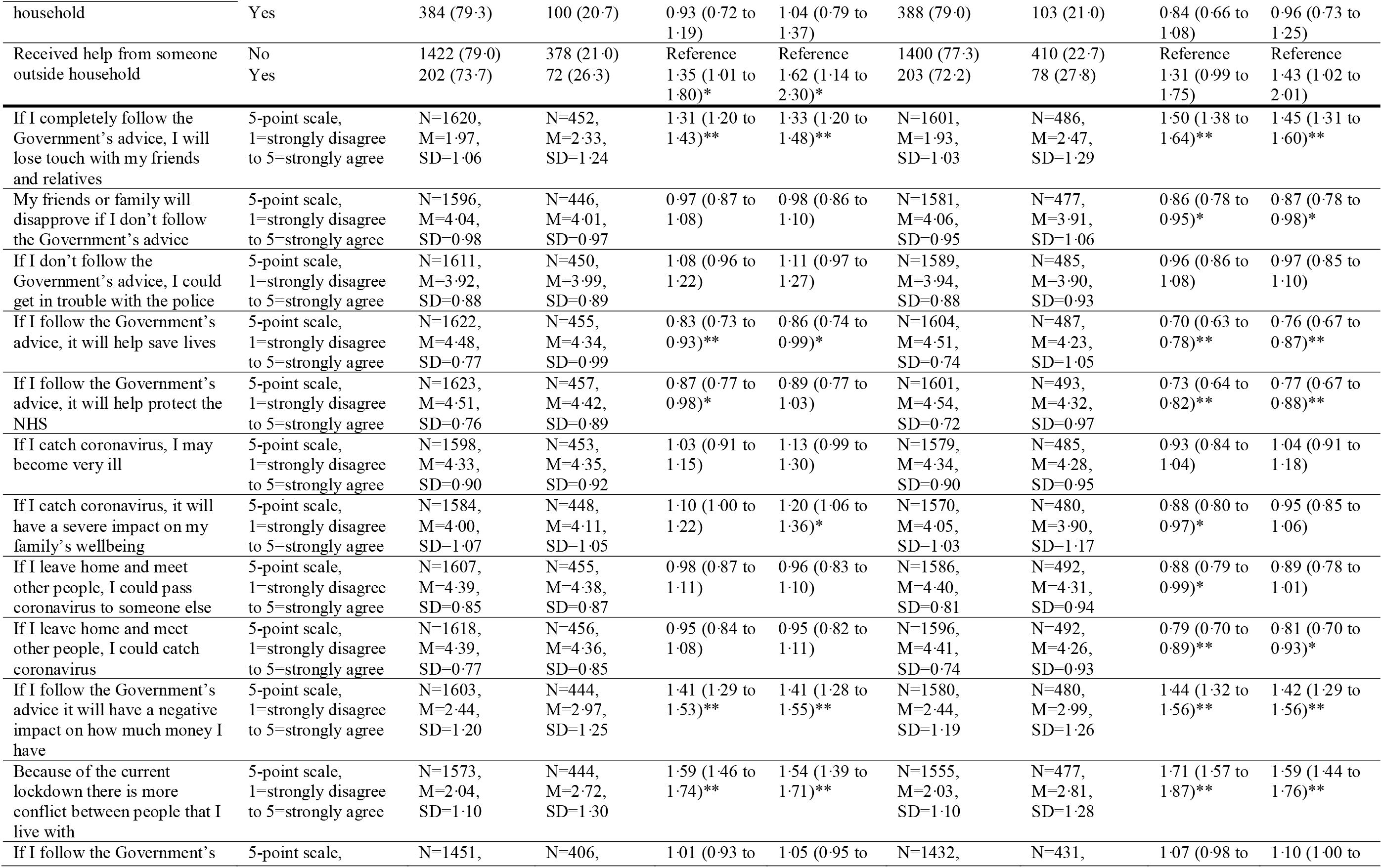
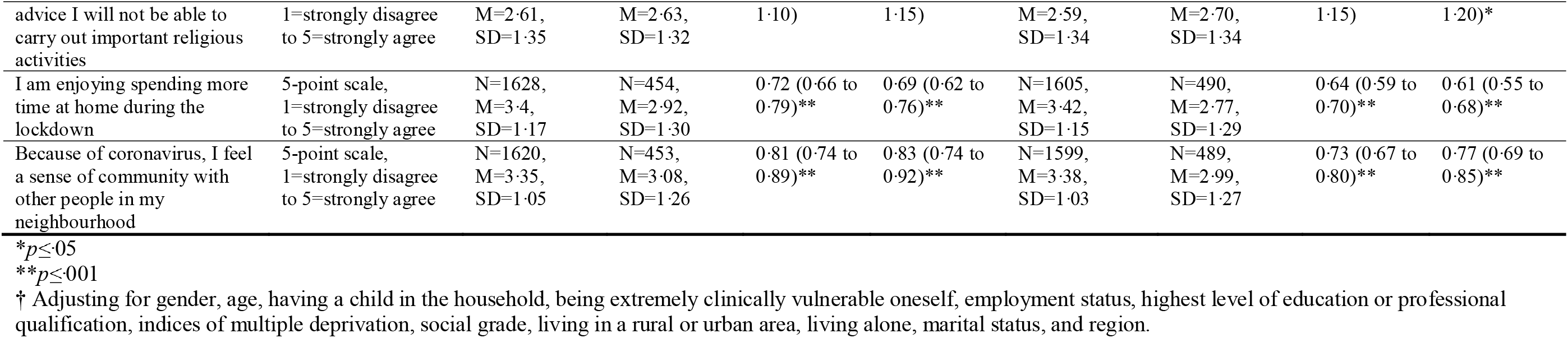
Associations between psychological and situational factors and anxiety and depression.

### Depression

Data were available to calculate depression scores for 2103 participants. 23·5% (n=494, 95% CI [21·7% to 25·3]) reported probable depression (scored three or over on PHQ-2).

Depression was associated with younger age, being clinically extremely vulnerable to COVID-19, not working, living in a more deprived area (Table 1). Those with a household member who was clinically extremely vulnerable to COVID-19 were less likely to report depression.

Depression was also associated with: presence of anxiety; feeling that the current lockdown had made your mental and physical health worse; presence of COVID-19 symptoms in the household; not enjoying spending more time at home because of the lockdown; more conflict in your household as a result of the current lockdown; believing that following Government advice would impact your finances negatively; cause you to lose touch with friends and relatives, and stop you from being able to carry out important religious activities; lower perceived effectiveness of Government measures; lower perceived sense of community with others in your neighbourhood; greater perceived likelihood of catching COVID-19; greater worry about COVID-19; greater perceived disapproval from family and friends if you did not follow Government advice; and thinking that fewer people of your age in the UK were following Government recommendations to stay at home (lower perceived social norms; Table 2).

### Self-reported general health

Self-reported general health was approximately normally distributed (N=2208, M=2·99, SD=1·04, median=3, mode=3).

Personal and clinical factors (gender, age, having a child in the household, being extremely clinically vulnerable oneself, employment status, highest level of education or professional qualification, indices of multiple deprivation, social grade, living in a rural or urban area, living alone, marital status, and region [results for region not reported]) explained 9·3% of the variance in self-reported general health (Table 3). Poorer self-reported general health was associated with being extremely clinically vulnerable to COVID-19 oneself, living in a more deprived area and lower social grade. Better self-reported general health was associated with being employed and being more educated (having a degree or higher).

**Table 3.**
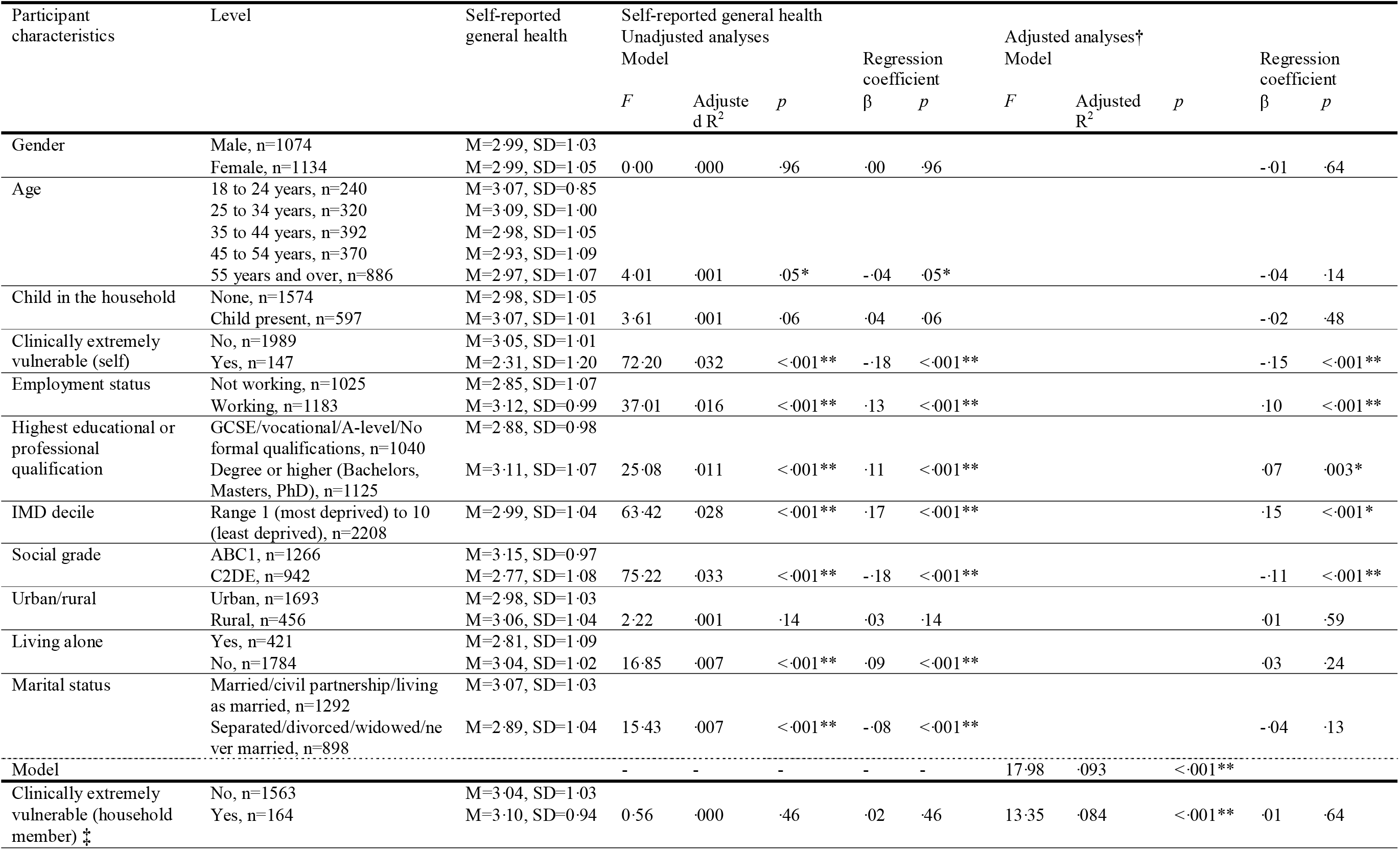
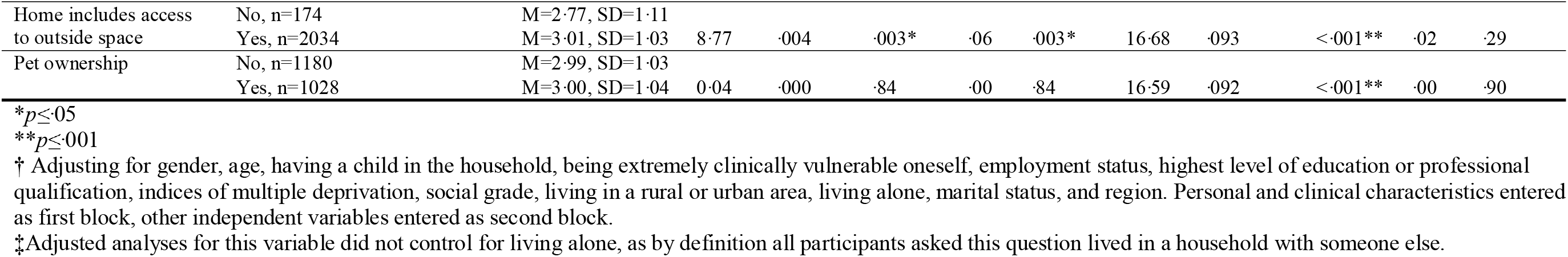
Associations between personal and clinical characteristics and self-reported general health.

Poorer self-reported general health was also associated with: thinking that the current lockdown had made your physical and mental health worse; presence of depression; presence of anxiety; reporting that you were currently self-isolating; greater perceived severity of COVID-19 (to self and family’s wellbeing); more conflict in your household as a result of the current lockdown; greater worry about COVID-19; presence of COVID-19 symptoms in the household; having received help from someone outside your household because of COVID-19; greater perceived likelihood of catching COVID-19; lower perceived effectiveness of Government measures; believing that following Government advice would impact your finances negatively; and thinking that you have had or currently have COVID-19 (Table 4). Factors associated with better self-reported general health were: greater total out of home activity; feeling an greater sense of community with people in your neighbourhood; thinking that more people of your age in the UK were following Government recommendations to stay at home (greater perceived social norms); enjoying spending more time at home during the lockdown; and having helped someone outside your household because of COVID-19 in the last week.

**Table 4.**
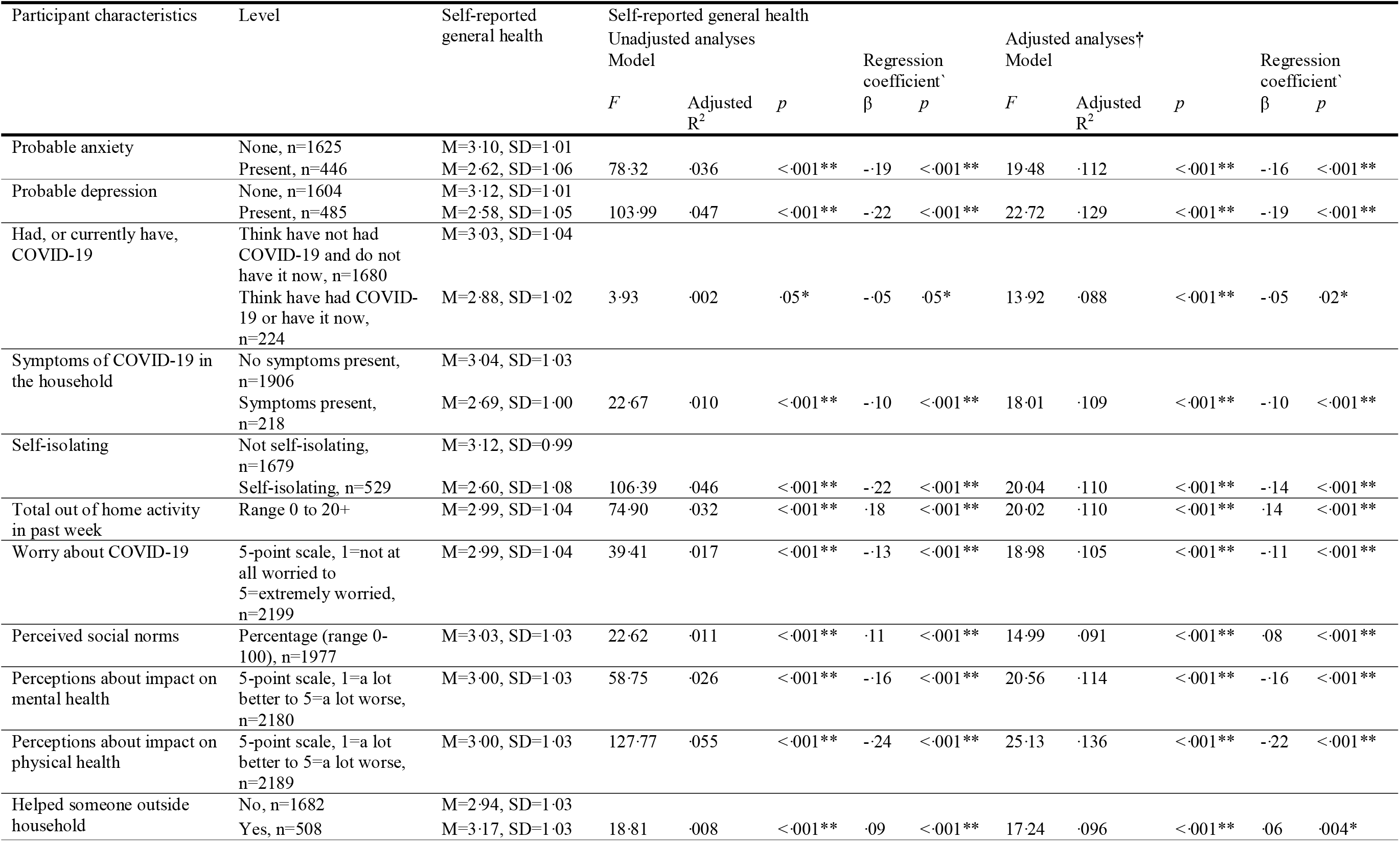
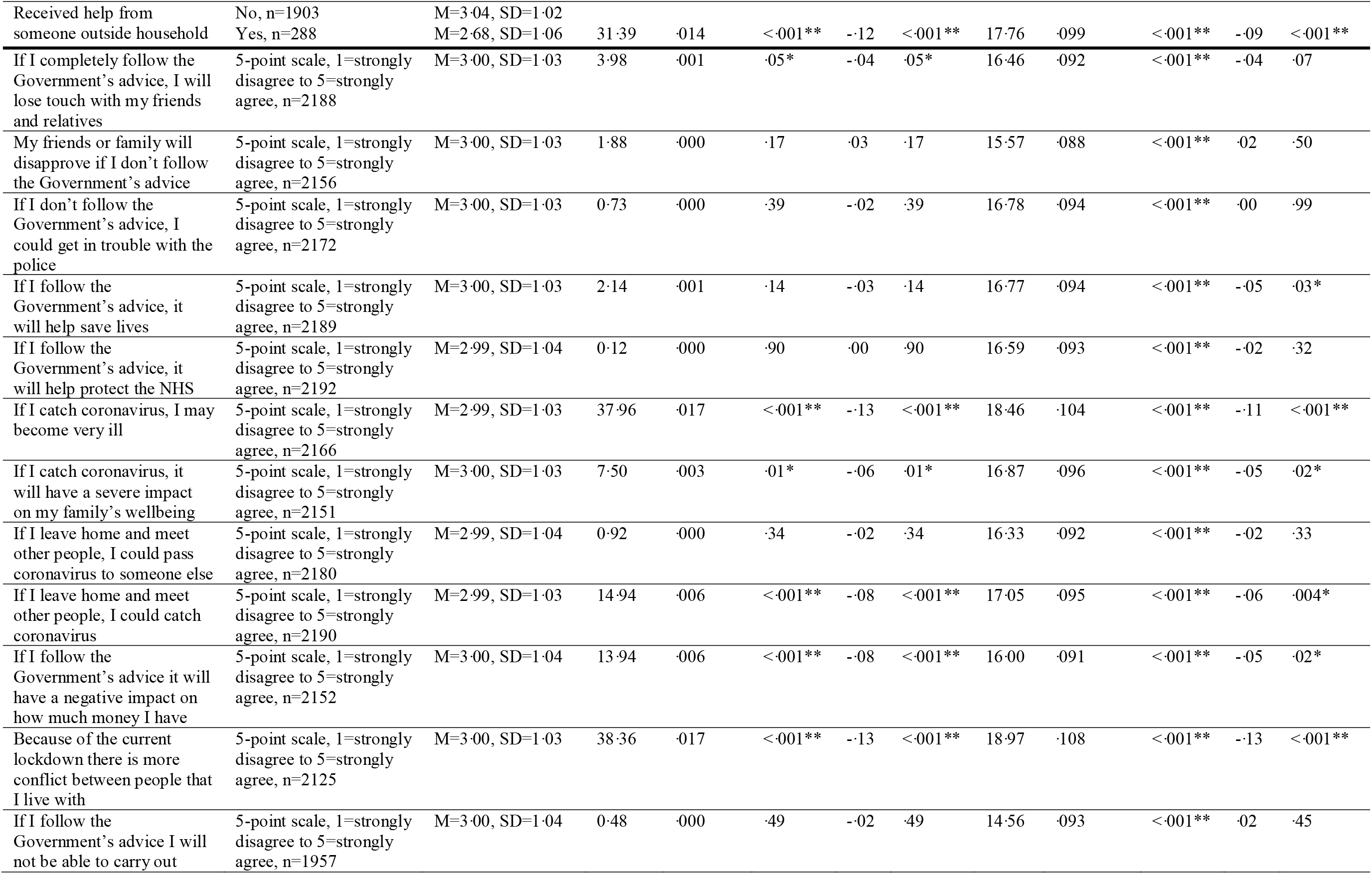
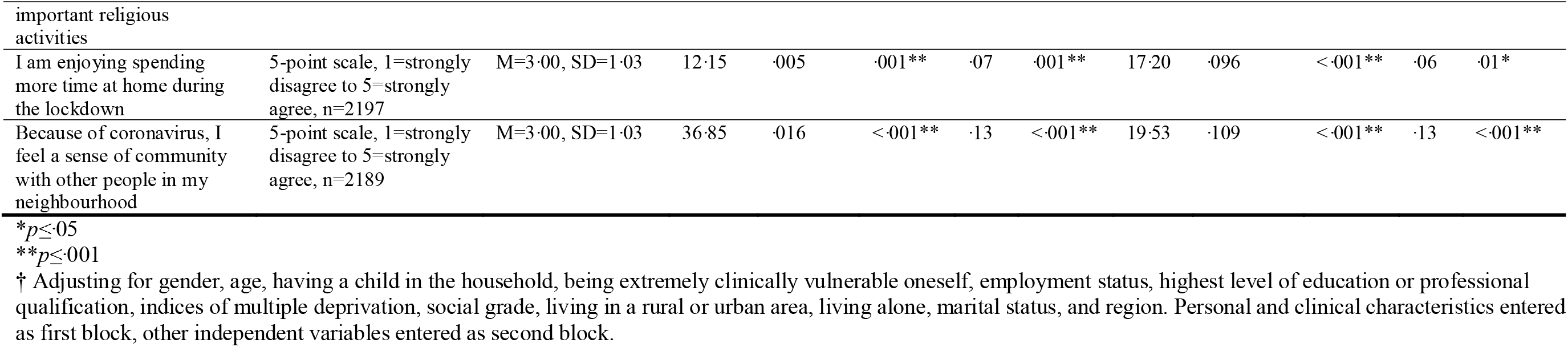
Associations between psychological and situational factors and self-reported general health.

## DISCUSSION

Reported rates of probable anxiety and depression in this sample were far higher than pre-lockdown population norms, with 21·9% reporting indicators compatible with clinical anxiety and 23·5% reporting indicators compatible with clinical depression. Normative data indicate that these rates are usually approximately 5% and 7%, respectively.^19^ The use of well-established measures that have normative data from the pre-pandemic period increases the methodological rigour of this study.^11^ As a point of comparison, using the same measure, the English national cohort study of flooding and health found only slightly higher levels of anxiety in their sample who had experienced flooding (28·3%) and slightly lower levels of depression (20·1%).^20^ While we cannot be certain about the absolute rates of anxiety and depression, high rates of psychological ill-health during lockdown have also been reported in other countries ^5–8^ as well as in the UK.^9,10^

Anxiety and depression were associated with markers of greater financial hardship, such as not working, living in a more deprived area, and thinking that if you followed the Government advice, it would have negative financial consequences.^1,10^ These findings suggest that tackling financial concerns may decrease psychological distress. Government measures should aim to provide extra support for those at greater risk of negative financial consequences. The negative financial impact of measures put in place to prevent the spread of COVID-19 has been greater among younger people and low-income households.^4^ Measures aiming to help with financial costs of the pandemic have been widely taken up in the UK, with 8.4 million workers having been “furloughed” through the UK Government job retention scheme (over one-quarter of all workers).^21^ However, those affected are likely to be concerned about uncertainties related to their job and future prospects. As the UK moves into the use of contact tracing to help prevent the spread of coronavirus as businesses and schools re-open, it is essential that people continue to be supported. This includes immediate reimbursement for the financial costs of self-isolation if they develop COVID-19 symptoms or are told that they have been in contact with someone who has COVID-19.^14^ While people isolating would be eligible for statutory sick pay if not entitled to a company-wide sick pay scheme,^22^ for many this would have a negative financial impact compared to their usual earnings.

Social factors were also associated with psychological distress in our sample. Anxiety and depression were associated with lower perceived social norms, thinking that you would lose touch with family or friends if you followed Government measures, greater conflict with the people you lived with, not enjoying time at home and lower sense of community with your neighbourhood. Other research has also found an association between depression and lower social support,^8^ and it is likely that limited support at least partly underlies our findings. However, the pattern of results also hints at a more corrosive cause, with conflict within households, declining solidarity with neighbours and feeling that others are not obeying the rules all impacting on mental health. As lockdown eases, fractures within communities may be appearing that are affecting morale and generating distress.

Being female and younger were also associated with a greater likelihood of anxiety and depression. Research in the UK and other countries during lockdown measures for COVID-19 has also found that females and those who are younger show greater levels of psychological distress.^5–9^ The prevalence of anxiety and depression is generally higher in females than males,^23^ and there is evidence that rates of anxiety and depression decrease with increasing age.^24^ Thus it is unsurprising that these characteristics are associated with anxiety and depression during the lockdown. However, findings from a series of cross-sectional surveys in the UK found that anxiety was higher in women than men in earlier stages of the lockdown, but that this gap narrowed over time.^10^ Evidence suggests that the burden of lockdown measures, especially in families with children, is differentially placed on females. A recent survey of over 3,500 families in the UK found that mothers were spending more time on household responsibilities than fathers, and were more likely to have lost their job, quit their job, or been furloughed.^25^ As schools begin to re-open in England, the burden of increased childcare may begin to ease for some parents, despite controversy around schools re-opening.^26^ Results from this study suggest that targeted communications and increased provision of support for those at greater risk of psychological distress may be warranted.

There was little evidence that illness-related factors, such as thinking that you have had or currently have COVID-19, presence of COVID-19 symptoms in the household, or greater perceived susceptibility of catching COVID-19 were associated with psychological distress. This is reassuring given the large number of people who have caught COVID-19 in the UK,^27^ and who may contract it in the future. However, lower perceived effectiveness of Government measures in preventing the spread of COVID-19 was associated with anxiety and depression. Results suggest that stressors associated with measures put in place to prevent the spread of COVID-19 were associated with psychological distress, rather than stressors associated with the illness itself. Greater worry about COVID-19 was also associated with psychological distress (note that there was no longer an association with depression when correcting for multiple comparisons).^9^ While greater worry is associated with increased adherence to lockdown measures,^14^ communications that decrease worry may also promote behaviours such as return to work.^28^ Highlighting that measures are effective is likely to decrease psychological distress, as well as promoting adherence.^14^

Poorer self-reported general health was associated with markers of greater financial hardship, such as living in a more deprived area, lower social grade, and having a chronic illness that makes you clinically extremely vulnerable to severe complications from COVID-19. Similarly, poorer psychological health was associated with poorer self-reported general health. These findings are in line with epidemiological findings indicating that greater inequality is associated with poorer health and wellbeing.^29^ Associations between psychological and situational factors and self-reported general health are difficult to disentangle given the cross-sectional nature of the study. However, results suggest plausible findings: those who reported that they were generally more healthy were more likely to have helped others due to COVID-19, while those who report that they are generally less healthy were more likely to have received help and to report that they were self-isolating.

This study has several limitations. First, while quota sampling was used and data were weighted to increase representativeness of the sample to the UK general population, we cannot be certain that views of survey respondents are representative of views of the general population.^30,31^ However, associations within the data still provide useful insights.^12^ Second, the cross-sectional nature of this study means that we are unable to imply causality. Third, we investigated self-reported general health using the first item of the SF-36, rather than the complete measure. Therefore, we were not able to compare data to population norms.^32^ This decision was taken due to time and space restrictions for the survey. Fourth, we calculated rates of probable anxiety and depression based on scores from self-report scales (GAD-2 and PHQ-2 respectively). However, these scales have been validated and are widely used as screening tools for psychological ill-health.^16,17^ Fifth, we did not measure ethnicity and so cannot say if mental health was differentially affected in different ethnic groups.

While we cannot be certain of the exact prevalence, rates of probable anxiety and depression in the UK were substantially higher during the lockdown than population norms. Most people rated their general health as “good”. Data suggest that psychological distress was associated with greater financial hardship, lower social support and greater conflict. Psychological distress was also associated with sociodemographic factors, such as being female, younger, unemployed, and living in a more deprived area. Poorer self-reported general health was also associated with greater inequality. These findings highlight the urgent need for better social and financial support for isolated, financially insecure young people to reduce the mental health impact of the pandemic and for greater attention among public health policy makers to the value of familial and community solidarity as potentially protective for mental health.

## Data Availability

Anonymised data will be made available upon reasonable request.

## DATA SHARING STATEMENT

Anonymised data will be made available upon reasonable request.

